# Criteria-based curation of a therapy-focused compendium to support treatment recommendations in precision oncology

**DOI:** 10.1101/2020.12.18.20248521

**Authors:** Frank P. Lin, Subotheni Thavaneswaran, John P. Grady, Mandy Ballinger, Maya Kansara, Samantha R. Oakes, Jayesh Desai, Chee Khoon Lee, John Simes, David M. Thomas

## Abstract

**BACKGROUND:** While several key resources exist that interpret therapeutic significance of genomic alterations in cancer, many regional real-world issues limit access to drugs. There is a need for a pragmatic, evidence-based, context-adapted tool to guide clinical management based on molecular biomarkers.

**METHODS:** A compendium of approved and experimental therapies with associated biomarkers was built following a survey of drug regulatory databases, existing knowledge bases, and published literature. Each biomarker-disease-therapy triplet was then categorized using a tiering system reflective of key therapeutic considerations: approved and reimbursed standard-of-care therapies with respect to a jurisdiction (Tier 1), evidence of efficacy or approval in another jurisdiction (Tier 2), evidence of antitumour activity (Tier 3), and plausible biological rationale (Tier 4). Two resistance categories were defined: lack of efficacy (Tier R1), and lack of antitumor activity (Tier R2).

**RESULTS:** Following comprehensive literature review and appraisal, we developed a curated knowledge base focused on drugs relevant and accessible in the Australian healthcare system (TOPOGRAPH: Therapy Oriented Precision Oncology Guidelines for Recommending Anticancer Pharmaceuticals). As of November 2020, TOPOGRAPH comprised 2810 biomarker-disease-therapy triplets in 989 expert-appraised entries, including 373 therapies, 199 predictive biomarkers, and 106 cancer types. In the 345 biomarker-linked therapies catalogued, 84 (24%) and 65 (19%) therapies in contexts of different cancer types have Tier 1 and 2 designations respectively, while 271 (79%) therapies were supported by preclinical studies, early clinical trials, retrospective studies, or case series (Tiers 3 and 4). A total of 119 of 373 (33%) therapies associated with biomarkers of resistance were also catalogued. A clinical algorithm was also developed to support therapeutic decision-making using predictive biomarkers. This resource is accessible online at https://topograph.info/.

**CONCLUSION:** TOPOGRAPH is intended to support oncologists with context-appropriate clinical decision-making– optimising selection and accessibility of the most appropriate targeted therapy for any given genomic biomarker. Our approach can be readily adapted to build jurisdiction-specific resources to standardise decision-making in precision oncology.

## Introduction

With tremendous progress in cancer biology and molecular diagnostics, cancer treatment is increasingly reliant on tumour molecular profiling to inform rational treatment decisions. To date, large precision oncology programs have harnessed the advances in genomic technology to inform design of a plethora of molecular matched trials [1-8]. Molecular testing, typically based on DNA and RNA sequencing, immunohistochemistry and *in situ* hybridization, is used to identify biomarkers that may predict response of cancers to particular targeted therapies. A number of uncontrolled trials have reported improved objective response rates and survival outcomes [7-11]. Outside the research setting, however, it is not clear how many patients benefit directly from genomically-informed therapies [12], highlighting the need for strategies to advance translation into clinical care.

A major challenge for oncologists is how best to integrate patients’ biomarker profiles into therapeutic decision-making [13,14]. The main barrier relates to the complex, rapidly evolving, and voluminous literature on the therapeutic significance of the detected biomarkers [15,16]. To aid clinicians, several knowledge bases have been developed by systematic cataloguing of molecular alterations in cancer to allow interpretation of variants and rapid interrogation of potential drug options [17-24]. The general concept of “actionability” – loosely defined as potential clinical utility of a biomarker for therapy selection [25] – is well-recognised and central to the value of a molecular assay. However, heterogeneous and often discordant metrics to assess evidence have created pervasive challenges [26], leading to efforts for consensus-based consolidation and harmonisation, to reduce variations in knowledge curation [26,27]

Variant-centred evidence taxonomies alone are insufficient to impact on clinical decisions based on molecular reports: oncologists ultimately make recommendations not only based on perceived efficacy of a therapy, but also considering potential harms and values relative to available options [28-30]. These complexities have prompted the development of more clinically-oriented criteria [22,25,31-32], which were recently harmonised into ESMO Scale for Clinical Actionability of molecular Targets (ESCAT) to standardise reporting of molecular recommendations [33].

Nonetheless, ESCAT has limitations in interpreting nuanced trial outcomes and specificities between drugs [34]. On the other hand, contextualised recommendations that build on local drug access is also critical for clinicians seeking therapeutic options for their patients. Specifically, availability of a drug plays a significant role in decision-making, but it varies considerably across different health systems; classification of a drug as “Food and Drug Administration (FDA)-approved” may not always be accessible outside the US, considering approval and reimbursement patterns.

To this end, we have built TOPOGRAPH - <u>T</u>herapy-<u>o</u>riented <u>p</u>recision <u>o</u>ncology <u>g</u>uidelines for recommendation of <u>a</u>nticancer <u>ph</u>armaceuticals – a dedicated, curated knowledge base that focuses on oncologists’ requirements in treatment selection based on contemporary biomarkers. A version specific to the Australian healthcare setting was developed to meet the clinical need of tiered evidence assessment for actionability, linking biomarkers to registered and experimental drugs, including through actively recruiting and accessible clinical trials. Through compilation of this compendium, we have established a curation process to enable consistency in evidence classification. We have further proposed a practical clinical algorithm to improve utilisation of this compendium, for rationalising therapeutic decision-making based on molecular biomarkers.

## Results

### Scope and objective of TOPOGRAPH

TOPOGRAPH established a catalogue of therapies – both approved/established and experimental – and their relationship with associated biomarkers and cancer types with respect to accessibility, efficacy, and antitumour activity (and lack thereof). Its objective is to provide a practical compendium to help oncologists with a context-adapted, prioritisation of treatment options in patients with advanced solid and haematological malignancies.

### Biomarkers curated by TOPOGRAPH

Following the curation process highlighted in Figure 1, TOPOGRAPH identified and catalogued biomarkers ranging from standard immunohistochemical assays (e.g., PD-L1 and oestrogen receptor) to molecular targets based on comprehensive genomic profiling. For genomic biomarkers, the curated biomarkers included genotypes of druggable molecular targets (e.g., mutations that sensitise a tyrosine kinase inhibitor or a therapeutic antibody), synthetically lethal candidates [e.g., *BRCA1/2* mutation and Poly (ADP-ribose) polymerase (PARP) inhibitors], as well as measurable signatures that are typically derived from a constellation of genomic changes (e.g., molecular phenotypes such as high TMB, microsatellite instability, and homologous recombination repair defects [35,36]) that demonstrably predict treatment outcomes.

**Figure 1.**
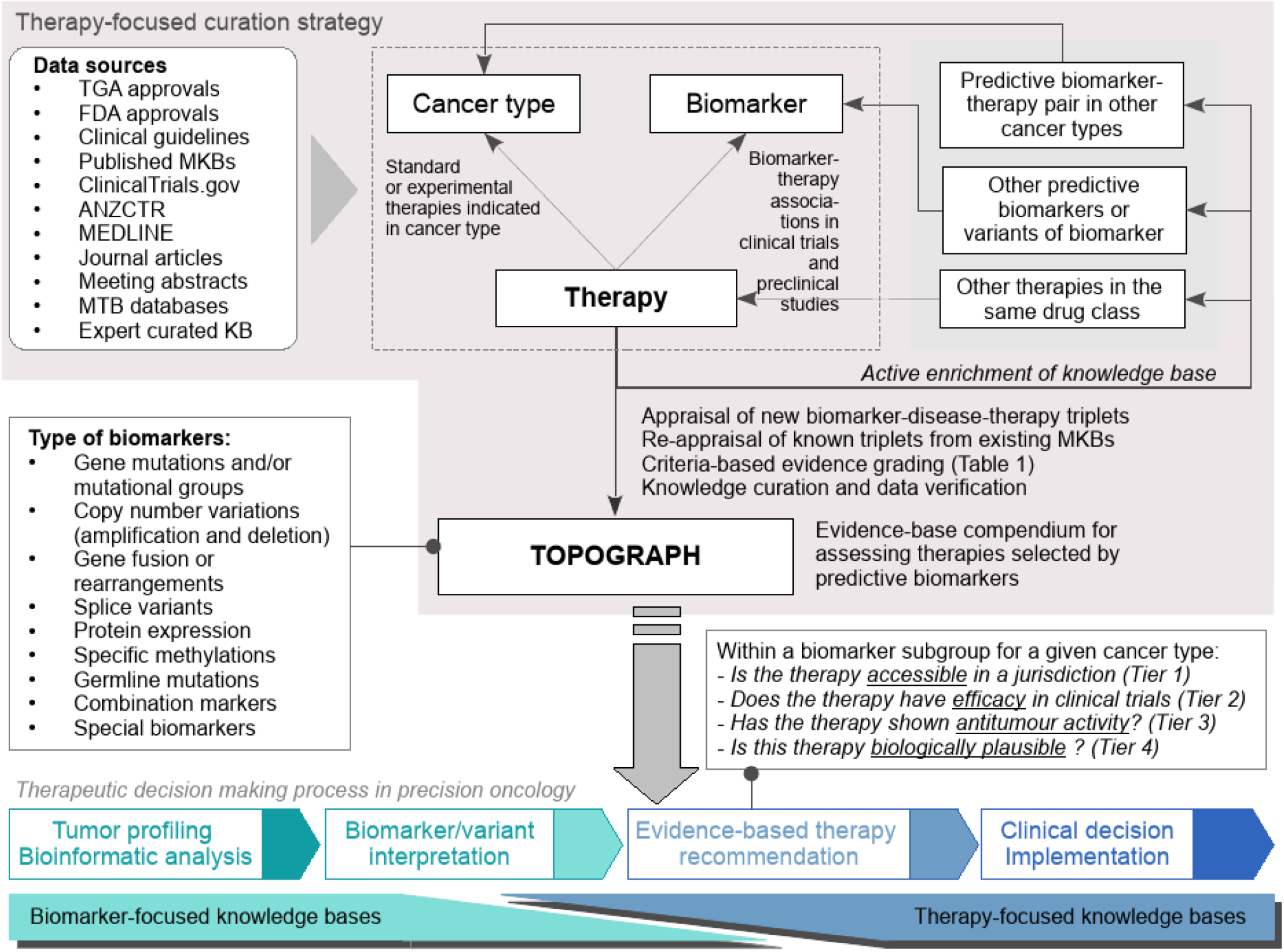
Method of data review and curation. The strategy used in curating TOPOGRAPH. Abbreviations: TGA: Therapeutic Goods Administration (Australia); FDA: Food and Drug Administration of United States; ANZCTR: Australia and New Zealand Clinical Trial Registry. KB: knowledge base.

At the time of data cut-off (November 2020), TOPOGRAPH contains 2810 triplets in 989 curated entries, comprising 887 unique alterations with potential therapeutic significance in 106 cancer types; the majority (N=814, 92%) being genomic targets potentially detectable by comprehensive genomic profiling (Table 2).

A total of 180 *variant groups* were defined to denote logical grouping of biomarkers with generalisable clinical significance [e.g., *EGFR* exon 19 deletions in non-small cell lung cancer (NSCLC)], including generic concepts of functional consequence of genetic alteration based on molecular pathology assessments (e.g, “oncogenic” or “gain-of-function” mutations). This compendium also enriched *combination biomarkers* (N=63), extending the definition of a biomarker beyond a single genotype, to emphasise their emerging roles in informing therapy (e.g., targeting both c-*MET* amplification in *EGFR*-mutated NSCLC has been studied as a strategy to overcome secondary resistance to gefitinib [37]). Similarly, *complex biomarkers* (N=6) were also catalogued to incorporate concepts of diagnostics that utilise signatures of underlying molecular processes to assist with therapy selection, (e.g., pembrolizumab in microsatellite instability-high or mismatch repair deficient colorectal cancer [38,39]).

### Therapy-focused criteria of evidence appraisal: tier designation

A literature-based criteria (Table 1 and Figure 2) was developed to systematically categorise information and evidence about accessibility, as well as efficacy, antitumour activities, and plausible biological rationale in the presence or absence of a given biomarker. As a general principle to rigorously assess the actionability of a biomarker, a higher tier was assigned if a biomarker was prospectively defined in the eligibility criteria of a clinical trial and the study was statistically powered to answer the biomarker question, indicating a greater strength of evidence. This is equivalent to categories A and B in Simon’s criteria with respect to the trial design, patient stratification, and statistical analysis elements [40]. Conversely, exploratory and retrospectively identified biomarkers, as well as biomarkers screened from observational studies, were considered hypothesis-generating and designated lower tiers (equivalent to Simon’s categories C & D).

**Table 1.**
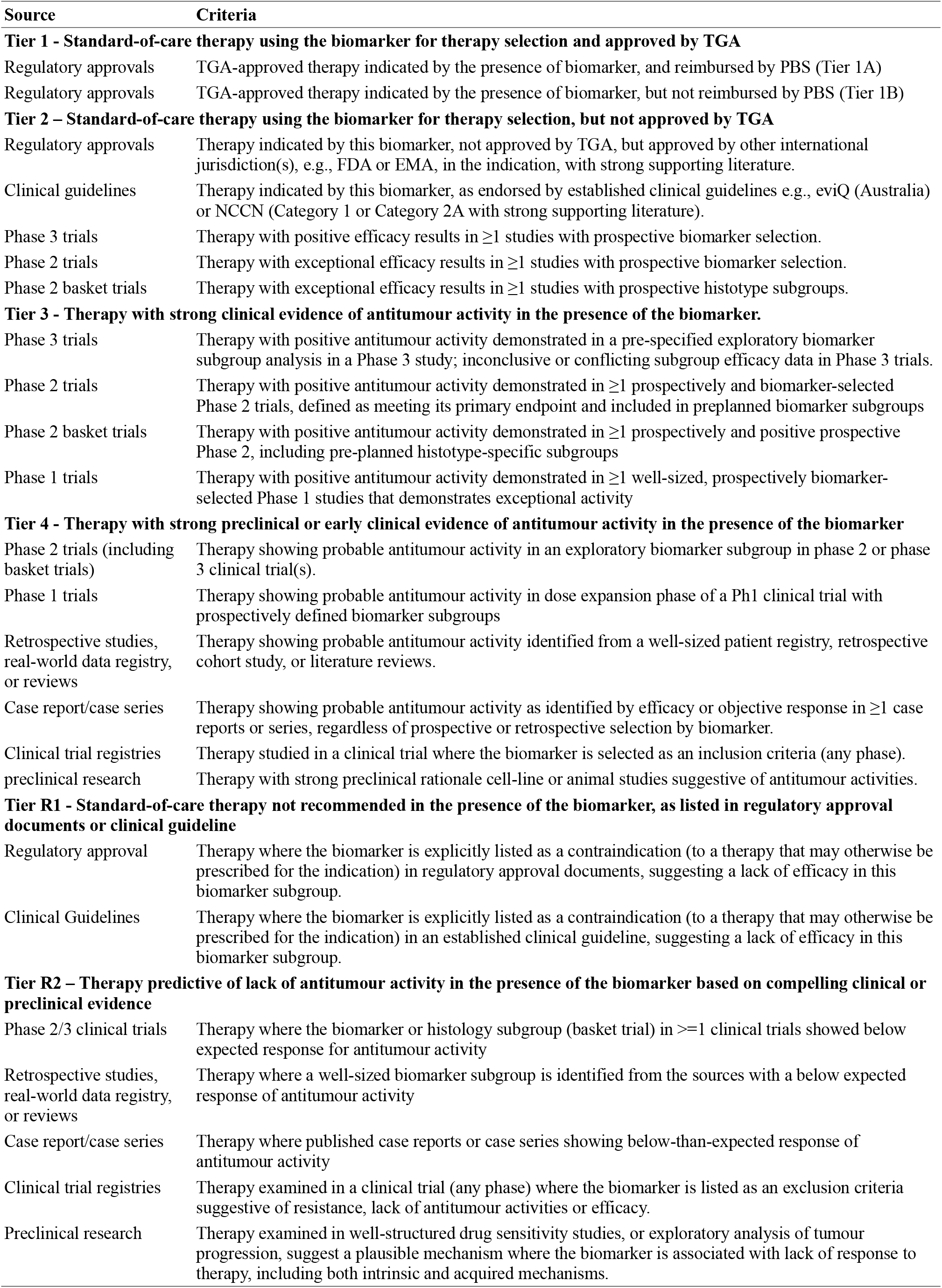
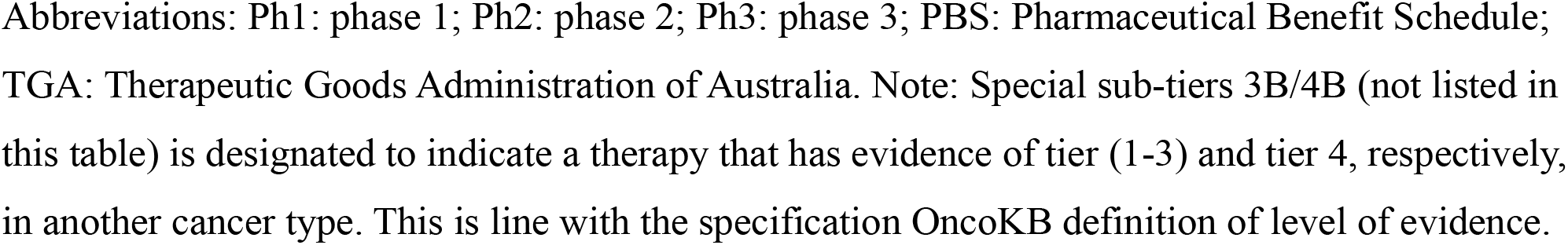
Definition of therapy recommendation tiers with literature assessment criteria used for curating TOPOGRAPH with therapies listed in Australia

| Source | Criteria |
| --- | --- |
| **Tier 1 - Standard-of-care therapy using the biomarker for therapy selection and approved by TGA** |  |
| Regulatory approvals | TGA-approved therapy indicated by the presence of biomarker, and reimbursed by PBS (Tier 1A) |
| Regulatory approvals | TGA-approved therapy indicated by the presence of biomarker, but not reimbursed by PBS (Tier 1B) |
| **Tier 2 – Standard-of-care therapy using the biomarker for therapy selection, but not approved by TGA** |  |
| Regulatory approvals | Therapy indicated by this biomarker, not approved by TGA, but approved by other international jurisdiction(s), e.g., FDA or EMA, in the indication, with strong supporting literature. |
| Clinical guidelines | Therapy indicated by this biomarker, as endorsed by established clinical guidelines e.g., eviQ (Australia) or NCCN (Category 1 or Category 2A with strong supporting literature). |
| Phase 3 trials | Therapy with positive efficacy results in $\geq 1$ studies with prospective biomarker selection. |
| Phase 2 trials | Therapy with exceptional efficacy results in $\geq 1$ studies with prospective biomarker selection. |
| Phase 2 basket trials | Therapy with exceptional efficacy results in $\geq 1$ studies with prospective histotype subgroups. |
| **Tier 3 - Therapy with strong clinical evidence of antitumour activity in the presence of the biomarker.** |  |
| Phase 3 trials | Therapy with positive antitumour activity demonstrated in a pre-specified exploratory biomarker subgroup analysis in a Phase 3 study; inconclusive or conflicting subgroup efficacy data in Phase 3 trials. |
| Phase 2 trials | Therapy with positive antitumour activity demonstrated in $\geq 1$ prospectively and biomarker-selected Phase 2 trials, defined as meeting its primary endpoint and included in preplanned biomarker subgroups |
| Phase 2 basket trials | Therapy with positive antitumour activity demonstrated in $\geq 1$ prospectively and positive prospective Phase 2, including pre-planned histotype-specific subgroups |
| Phase 1 trials | Therapy with positive antitumour activity demonstrated in $\geq 1$ well-sized, prospectively biomarker-selected Phase 1 studies that demonstrates exceptional activity |
| **Tier 4 - Therapy with strong preclinical or early clinical evidence of antitumour activity in the presence of the biomarker** |  |
| Phase 2 trials (including basket trials) | Therapy showing probable antitumour activity in an exploratory biomarker subgroup in phase 2 or phase 3 clinical trial(s). |
| Phase 1 trials | Therapy showing probable antitumour activity in dose expansion phase of a Ph1 clinical trial with prospectively defined biomarker subgroups |
| Retrospective studies, real-world data registry, or reviews | Therapy showing probable antitumour activity identified from a well-sized patient registry, retrospective cohort study, or literature reviews. |
| Case report/case series | Therapy showing probable antitumour activity as identified by efficacy or objective response in $\geq 1$ case reports or series, regardless of prospective or retrospective selection by biomarker. |
| Clinical trial registries | Therapy studied in a clinical trial where the biomarker is selected as an inclusion criteria (any phase). |
| preclinical research | Therapy with strong preclinical rationale cell-line or animal studies suggestive of antitumour activities. |
| **Tier R1 - Standard-of-care therapy not recommended in the presence of the biomarker, as listed in regulatory approval documents or clinical guideline** |  |
| Regulatory approval | Therapy where the biomarker is explicitly listed as a contraindication (to a therapy that may otherwise be prescribed for the indication) in regulatory approval documents, suggesting a lack of efficacy in this biomarker subgroup. |
| Clinical Guidelines | Therapy where the biomarker is explicitly listed as a contraindication (to a therapy that may otherwise be prescribed for the indication) in an established clinical guideline, suggesting a lack of efficacy in this biomarker subgroup. |
| **Tier R2 – Therapy predictive of lack of antitumour activity in the presence of the biomarker based on compelling clinical or preclinical evidence** |  |
| Phase 2/3 clinical trials | Therapy where the biomarker or histology subgroup (basket trial) in $\geq 1$ clinical trials showed below expected response for antitumour activity |
| Retrospective studies, real-world data registry, or reviews | Therapy where a well-sized biomarker subgroup is identified from the sources with a below expected response of antitumour activity |
| Case report/case series | Therapy where published case reports or case series showing below-than-expected response of antitumour activity |
| Clinical trial registries | Therapy examined in a clinical trial (any phase) where the biomarker is listed as an exclusion criteria suggestive of resistance, lack of antitumour activities or efficacy. |
| Preclinical research | Therapy examined in well-structured drug sensitivity studies, or exploratory analysis of tumour progression, suggest a plausible mechanism where the biomarker is associated with lack of response to therapy, including both intrinsic and acquired mechanisms. |

Abbreviations: Ph1: phase 1; Ph2: phase 2; Ph3: phase 3; PBS: Pharmaceutical Benefit Schedule; TGA: Therapeutic Goods Administration of Australia. Note: Special sub-tiers 3B/4B (not listed in this table) is designated to indicate a therapy that has evidence of tier (1-3) and tier 4, respectively, in another cancer type. This is line with the specification OncoKB definition of level of evidence.

**Table 2.**
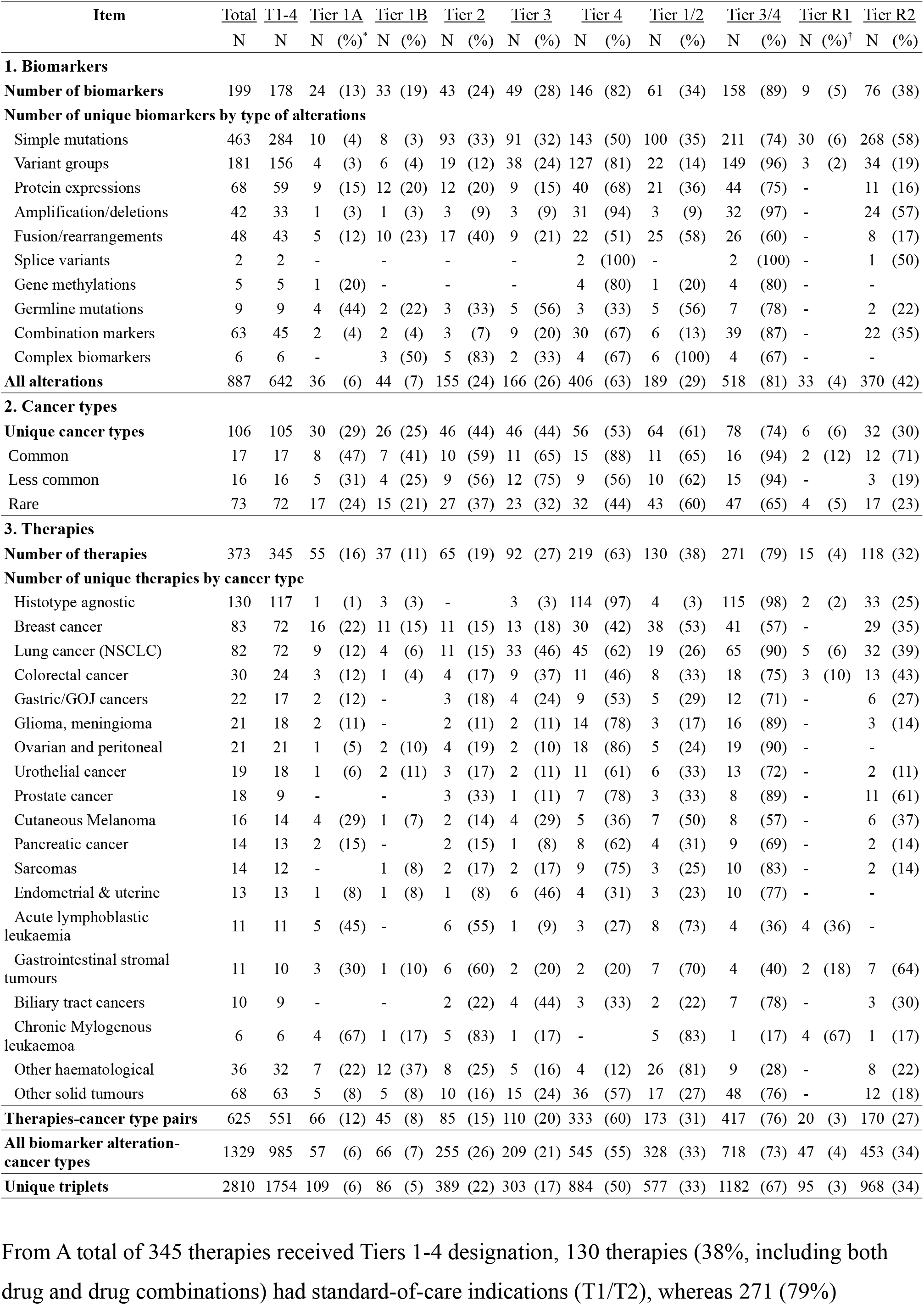
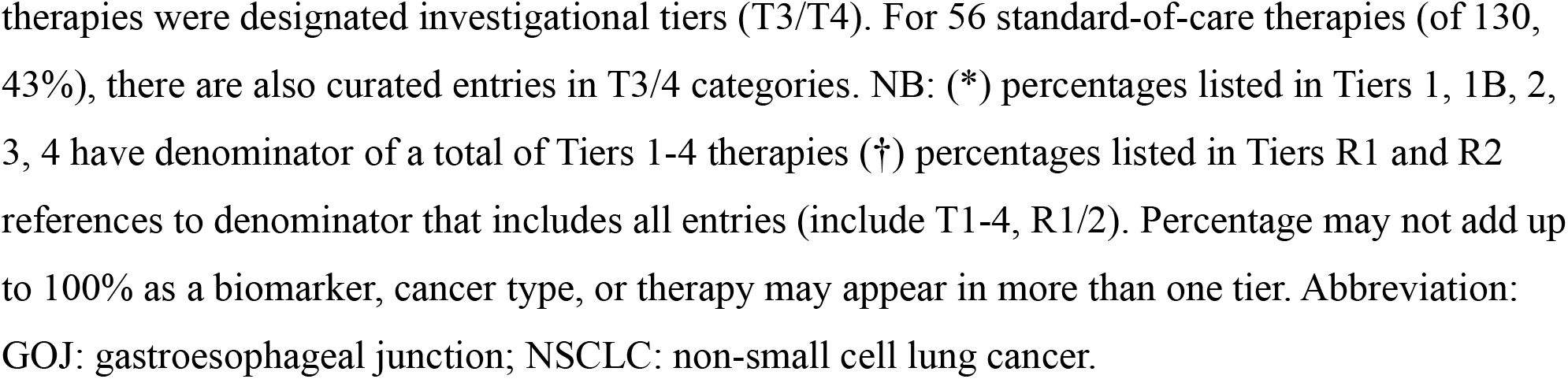
Summary of biomarkers and therapies curated in TOPOGRAPH by tiers

| Item | Total<br>N | T1-4<br>N | Tier 1A<br>N (%)<sup>*</sup> | Tier 1B<br>N (%) | Tier 2<br>N (%) | Tier 3<br>N (%) | Tier 4<br>N (%) | Tier 1/2<br>N (%) | Tier 3/4<br>N (%) | Tier R1<br>N (%)<sup>†</sup> | Tier R2<br>N (%) |
| --- | --- | --- | --- | --- | --- | --- | --- | --- | --- | --- | --- |
| **1. Biomarkers** |  |  |  |  |  |  |  |  |  |  |  |
| **Number of biomarkers** | 199 | 178 | 24 (13) | 33 (19) | 43 (24) | 49 (28) | 146 (82) | 61 (34) | 158 (89) | 9 (5) | 76 (38) |
| **Number of unique biomarkers by type of alterations** |  |  |  |  |  |  |  |  |  |  |  |
| Simple mutations | 463 | 284 | 10 (4) | 8 (3) | 93 (33) | 91 (32) | 143 (50) | 100 (35) | 211 (74) | 30 (6) | 268 (58) |
| Variant groups | 181 | 156 | 4 (3) | 6 (4) | 19 (12) | 38 (24) | 127 (81) | 22 (14) | 149 (96) | 3 (2) | 34 (19) |
| Protein expressions | 68 | 59 | 9 (15) | 12 (20) | 12 (20) | 9 (15) | 40 (68) | 21 (36) | 44 (75) | - | 11 (16) |
| Amplification/deletions | 42 | 33 | 1 (3) | 1 (3) | 3 (9) | 3 (9) | 31 (94) | 3 (9) | 32 (97) | - | 24 (57) |
| Fusion/rearrangements | 48 | 43 | 5 (12) | 10 (23) | 17 (40) | 9 (21) | 22 (51) | 25 (58) | 26 (60) | - | 8 (17) |
| Splice variants | 2 | 2 | - | - | - | - | 2 (100) | - | 2 (100) | - | 1 (50) |
| Gene methylations | 5 | 5 | 1 (20) | - | - | - | 4 (80) | 1 (20) | 4 (80) | - | - |
| Germline mutations | 9 | 9 | 4 (44) | 2 (22) | 3 (33) | 5 (56) | 3 (33) | 5 (56) | 7 (78) | - | 2 (22) |
| Combination markers | 63 | 45 | 2 (4) | 2 (4) | 3 (7) | 9 (20) | 30 (67) | 6 (13) | 39 (87) | - | 22 (35) |
| Complex biomarkers | 6 | 6 | - | 3 (50) | 5 (83) | 2 (33) | 4 (67) | 6 (100) | 4 (67) | - | - |
| **All alterations** | 887 | 642 | 36 (6) | 44 (7) | 155 (24) | 166 (26) | 406 (63) | 189 (29) | 518 (81) | 33 (4) | 370 (42) |
| **2. Cancer types** |  |  |  |  |  |  |  |  |  |  |  |
| **Unique cancer types** | 106 | 105 | 30 (29) | 26 (25) | 46 (44) | 46 (44) | 56 (53) | 64 (61) | 78 (74) | 6 (6) | 32 (30) |
| Common | 17 | 17 | 8 (47) | 7 (41) | 10 (59) | 11 (65) | 15 (88) | 11 (65) | 16 (94) | 2 (12) | 12 (71) |
| Less common | 16 | 16 | 5 (31) | 4 (25) | 9 (56) | 12 (75) | 9 (56) | 10 (62) | 15 (94) | - | 3 (19) |
| Rare | 73 | 72 | 17 (24) | 15 (21) | 27 (37) | 23 (32) | 32 (44) | 43 (60) | 47 (65) | 4 (5) | 17 (23) |
| **3. Therapies** |  |  |  |  |  |  |  |  |  |  |  |
| **Number of therapies** | 373 | 345 | 55 (16) | 37 (11) | 65 (19) | 92 (27) | 219 (63) | 130 (38) | 271 (79) | 15 (4) | 118 (32) |
| **Number of unique therapies by cancer type** |  |  |  |  |  |  |  |  |  |  |  |
| Histotype agnostic | 130 | 117 | 1 (1) | 3 (3) | - | 3 (3) | 114 (97) | 4 (3) | 115 (98) | 2 (2) | 33 (25) |
| Breast cancer | 83 | 72 | 16 (22) | 11 (15) | 11 (15) | 13 (18) | 30 (42) | 38 (53) | 41 (57) | - | 29 (35) |
| Lung cancer (NSCLC) | 82 | 72 | 9 (12) | 4 (6) | 11 (15) | 33 (46) | 45 (62) | 19 (26) | 65 (90) | 5 (6) | 32 (39) |
| Colorectal cancer | 30 | 24 | 3 (12) | 1 (4) | 4 (17) | 9 (37) | 11 (46) | 8 (33) | 18 (75) | 3 (10) | 13 (43) |
| Gastric/GOJ cancers | 22 | 17 | 2 (12) | - | 3 (18) | 4 (24) | 9 (53) | 5 (29) | 12 (71) | - | 6 (27) |
| Glioma, meningioma | 21 | 18 | 2 (11) | - | 2 (11) | 2 (11) | 14 (78) | 3 (17) | 16 (89) | - | 3 (14) |
| Ovarian and peritoneal | 21 | 21 | 1 (5) | 2 (10) | 4 (19) | 2 (10) | 18 (86) | 5 (24) | 19 (90) | - | - |
| Urothelial cancer | 19 | 18 | 1 (6) | 2 (11) | 3 (17) | 2 (11) | 11 (61) | 6 (33) | 13 (72) | - | 2 (11) |
| Prostate cancer | 18 | 9 | - | - | 3 (33) | 1 (11) | 7 (78) | 3 (33) | 8 (89) | - | 11 (61) |
| Cutaneous Melanoma | 16 | 14 | 4 (29) | 1 (7) | 2 (14) | 4 (29) | 5 (36) | 7 (50) | 8 (57) | - | 6 (37) |
| Pancreatic cancer | 14 | 13 | 2 (15) | - | 2 (15) | 1 (8) | 8 (62) | 4 (31) | 9 (69) | - | 2 (14) |
| Sarcomas | 14 | 12 | - | 1 (8) | 2 (17) | 2 (17) | 9 (75) | 3 (25) | 10 (83) | - | 2 (14) |
| Endometrial & uterine | 13 | 13 | 1 (8) | 1 (8) | 1 (8) | 6 (46) | 4 (31) | 3 (23) | 10 (77) | - | - |
| Acute lymphoblastic leukaemia | 11 | 11 | 5 (45) | - | 6 (55) | 1 (9) | 3 (27) | 8 (73) | 4 (36) | 4 (36) | - |
| Gastrointestinal stromal tumours | 11 | 10 | 3 (30) | 1 (10) | 6 (60) | 2 (20) | 2 (20) | 7 (70) | 4 (40) | 2 (18) | 7 (64) |
| Biliary tract cancers | 10 | 9 | - | - | 2 (22) | 4 (44) | 3 (33) | 2 (22) | 7 (78) | - | 3 (30) |
| Chronic Myelogenous leukaemia | 6 | 6 | 4 (67) | 1 (17) | 5 (83) | 1 (17) | - | 5 (83) | 1 (17) | 4 (67) | 1 (17) |
| Other haematological | 36 | 32 | 7 (22) | 12 (37) | 8 (25) | 5 (16) | 4 (12) | 26 (81) | 9 (28) | - | 8 (22) |
| Other solid tumours | 68 | 63 | 5 (8) | 5 (8) | 10 (16) | 15 (24) | 36 (57) | 17 (27) | 48 (76) | - | 12 (18) |
| **Therapies-cancer type pairs** | 625 | 551 | 66 (12) | 45 (8) | 85 (15) | 110 (20) | 333 (60) | 173 (31) | 417 (76) | 20 (3) | 170 (27) |
| **All biomarker alteration-cancer types** | 1329 | 985 | 57 (6) | 66 (7) | 255 (26) | 209 (21) | 545 (55) | 328 (33) | 718 (73) | 47 (4) | 453 (34) |
| **Unique triplets** | 2810 | 1754 | 109 (6) | 86 (5) | 389 (22) | 303 (17) | 884 (50) | 577 (33) | 1182 (67) | 95 (3) | 968 (34) |

From A total of 345 therapies received Tiers 1-4 designation, 130 therapies (38%, including both drug and drug combinations) had standard-of-care indications (T1/T2), whereas 271 (79%)
therapies were designated investigational tiers (T3/T4). For 56 standard-of-care therapies (of 130, 43%), there are also curated entries in T3/4 categories. NB: (\*) percentages listed in Tiers 1, 1B, 2, 3, 4 have denominator of a total of Tiers 1-4 therapies (†) percentages listed in Tiers R1 and R2 references to denominator that includes all entries (include T1-4, R1/2). Percentage may not add up to 100% as a biomarker, cancer type, or therapy may appear in more than one tier. Abbreviation: GOJ: gastroesophageal junction; NSCLC: non-small cell lung cancer.

**Figure 2.**
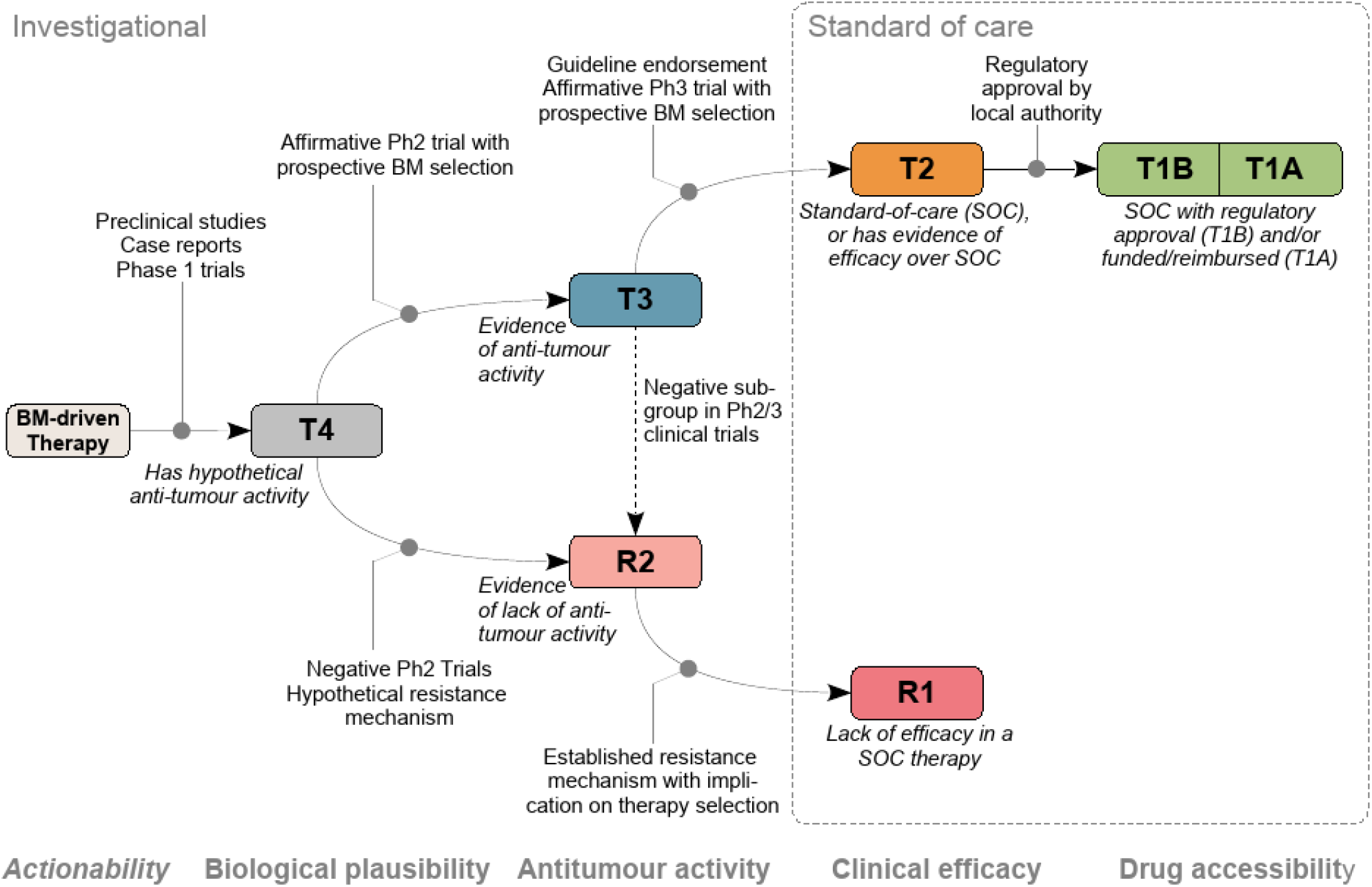
Hierarchy of tiers curated in TOPOGRAPH. The definition of evidence tiers in TOPOGRAPH reflects the maturity of a drug in its development. Tiers T1, T2, and R1 are considered standard-of-care, whereas the remaining tiers are investigational. Therapy yet to be adequately studied for possible clinical activities are designated T4. Abbreviation: BM: Biomarker; Ph2: phase 2 clinical trials; Ph3: Phase 3 clinical trials; SOC standard of care.

Therapies designated *Tier 1* (T1) are drugs (including combinations) approved by the local regulatory authority and indicated for treatment in the context of the relevant cancer type and biomarker. The definition is similar to top-level categories in clinical classification schemes [22,25,31]. Through reviewing the list of approved cancer drugs in Australia, a total 84 (24%) of 373 therapies have a biomarker mandated as a part of therapy indications; extracted from the Therapeutic Goods Administration (TGA, equivalent authority to FDA). Considering the importance of financial burden on patients when accessing a drug, T1 therapies were further dichotomised into 1A and 1B to reflect funding status of a drug within the healthcare system. As of November 2020, 55 therapies were publicly reimbursed in Australia through the Pharmaceutical Benefit Scheme (Tier 1A, 16%) [41]. Thirty-seven therapies (Tier 1B, 11%) were approved by TGA but not publicly funded.

*Tier 2* (T2) therapies have proven efficacies in large clinical trials where the prospective biomarker is used for treatment selection, but is distinguished from T1 given its lack of jurisdiction approval. Therapies approved in other jurisdictions are found in this tier designation. For example, FDA-approved drugs and standard-of-care therapies endorsed by major guidelines – but not approved by the TGA - are included in Tier 2, as these drugs may have relevance to clinical decision-making.

Rarely, therapies in early-phase trials that have demonstrated exceptional efficacy may warrant being catalogued. This category is important because the cost of therapy may result in out-of-pocket expenses, necessitating access via compassionate, or special access programs, given that they are commercially available elsewhere. A total of 65 therapies (19%) were designated T2 in TOPOGRAPH at data cut-off.

Most T1 and T2 therapies are assigned based on the actual or potential drug approvals, although critical review of results of clinical trials is needed to scrutinise mixed efficacy within composite predictive biomarker or histotype subgroups. For example, in a phase 2 basket trial of pembrolizumab in advanced cancers (KEYNOTE-158), the use of TMB as a companion biomarker for pembrolizumab has seen different treatment responses across histotypes [42], leading to expert panels calling for more judicious testing of TMB only in certain cancer types [43]. Heterogeneous outcomes within subgroups of clinical trial are captured and differentially tiered in TOPOGRAPH (Supplementary Table S1).

*Tier 3* (T3) therapies have demonstrated clinical activity in the presence of the biomarker, based on a substantive phase 2 trial or equivalent (Table 1). Considering heterogeneous outcomes are used in different trials, a “positive” result is defined as one that meets its predefined primary endpoint, including one or more prospectively specified biomarker subgroups as part of the inclusion criteria. The definition of T3 is largely equivalent to the Tier II category in ESCAT, highlighting therapies yet to have proven efficacy in larger trials. As described above, we have de-emphasised the retrospectively defined biomarkers to lower tiers, given that influences from confounding factors cannot be conclusively ascertained. A total of 92 (27%) were curated as T3 in TOPOGRAPH at data cut-off.

*Tier 4* (T4) therapies in the presence of a biomarker are considered hypothesis generating. T4 therapies include all retrospectively identified or exploratory biomarkers associated with antitumour activity. This tier is largely concordant to ESCAT levels III and IV. The types of evidence that support the designation of T4 include case reports, preclinical cell-line and animal studies, and plausible signals identified from real-world evidence. T4 therapies are important in drug development, such that referral to early phase clinical trials is usually encouraged in the absence of other therapy options with a higher tier. As such, therapies being studied in a clinical trial that specifically include the biomarker as an eligibility criterion were also candidates for curation. More than half of curated therapies have T4 evidence entries (N=219, 63%).

Therapies designated *R1* have evidence to suggest lack of efficacy in the presence of the biomarker, and these treatments are often explicitly *not* recommended by the regulatory authorities where the therapy would otherwise be indicated. In effect, the presence of R1 biomarker negates evidence for efficacy otherwise indicated by T1/2 tier. The definition of R1 is largely concordant to the corresponding LOE in OncoKB (e.g., *KRAS* exons 2-4 mutations for anti-EGFR antibodies in advanced colorectal cancers).

Therapies designated *R2* have evidence to show lack of clinical or preclinical activity in the presence of the biomarker, typically from phase 2 studies that did not meet their predefined biomarker-selected primary endpoints (Figure 2). Cataloguing R2 entries is helpful in prioritising treatment decisions: in situations where a therapy would otherwise be rationally recommended, the presence of a biomarker in this category may reduce the overall strength-of-recommendation of a treatment. We supplemented our literature criteria for R2 by inclusion of preclinical or pharmacodynamic studies that compare half-maximal inhibitory concentrations (IC50) of different targetable mutations as a proxy for likely clinical response. Since markers of drug resistance are difficult to study prospectively, such information provides a useful source of additional information to guide decision-making. Observational studies also contribute to a significant body of knowledge in this tier, as well as studies that use paired before-after tumour profiling to identify resistance mechanisms following therapy exposure. Of note, the active enrichment process has significantly increased the volume of curated triplets: a total number of simple mutations across all biomarkers designated R2 (268, 58% of all mutations), with the majority from preclinical evidence.

### Patterns of biomarker-selected therapies and their associations with cancer types

Across all cancer types, non-small cell lung (N=72) and breast cancers (N=72) had the most targeted therapies and/or combination therapy options. Cancer types with more than half of curated targeted therapy available as a standard care included chronic myelogenous leukaemia (5 of 6 curated, 83%), acute lymphoblastic leukaemia (N=8, 73%), gastrointestinal stromal tumours (GIST, N=7, 70%), cutaneous melanoma (N=7, 50%), and breast cancer (N=38, 53%). Four standard-of-care therapies (3%) were indicated solely on biomarkers without specific reference to histology (T1/T2).

We further hypothesised that histotype-specific evidence may disadvantage rare cancer populations. To examine this, we further classified 106 curated cancer types into common (incidence of ≥12 per 100,000 person-year, N=17), less common (≥6 and <12 per 100,000, N=16), and rare subgroups (<6 per 100,000, N=73). For standard-of-care therapies, 11 common (65%), 10 less common (62%), and 43 rare (60%) cancer types have at least one T1/2 therapy curated in the TOPOGRAPH database (p=0.89, Pearson’s Chi-square test with 2 degrees of freedom). Conversely, while almost all common and less common cancer types have experimental therapies (i.e., T3/4; common: 16, 94% and less common: 15, 94%), only 47 rare cancers (65%) have histotype-specific, biomarker-selected therapy under investigation (p=0.0061, Chi-square test with 2 d.f.).

### Proposal of a decision algorithm for rationalising biomarker-driven therapy recommendation

To standardise therapy recommendations based on biomarkers, we propose a cascading decision algorithm to rationalise prioritisation of potential treatment options (Figure 3). The proposed algorithm is largely concordant with recommendations proposed in the ESCAT criteria [33], although there is a strong emphasis on clinical trial participation in our recommendations.

**Figure 3.**
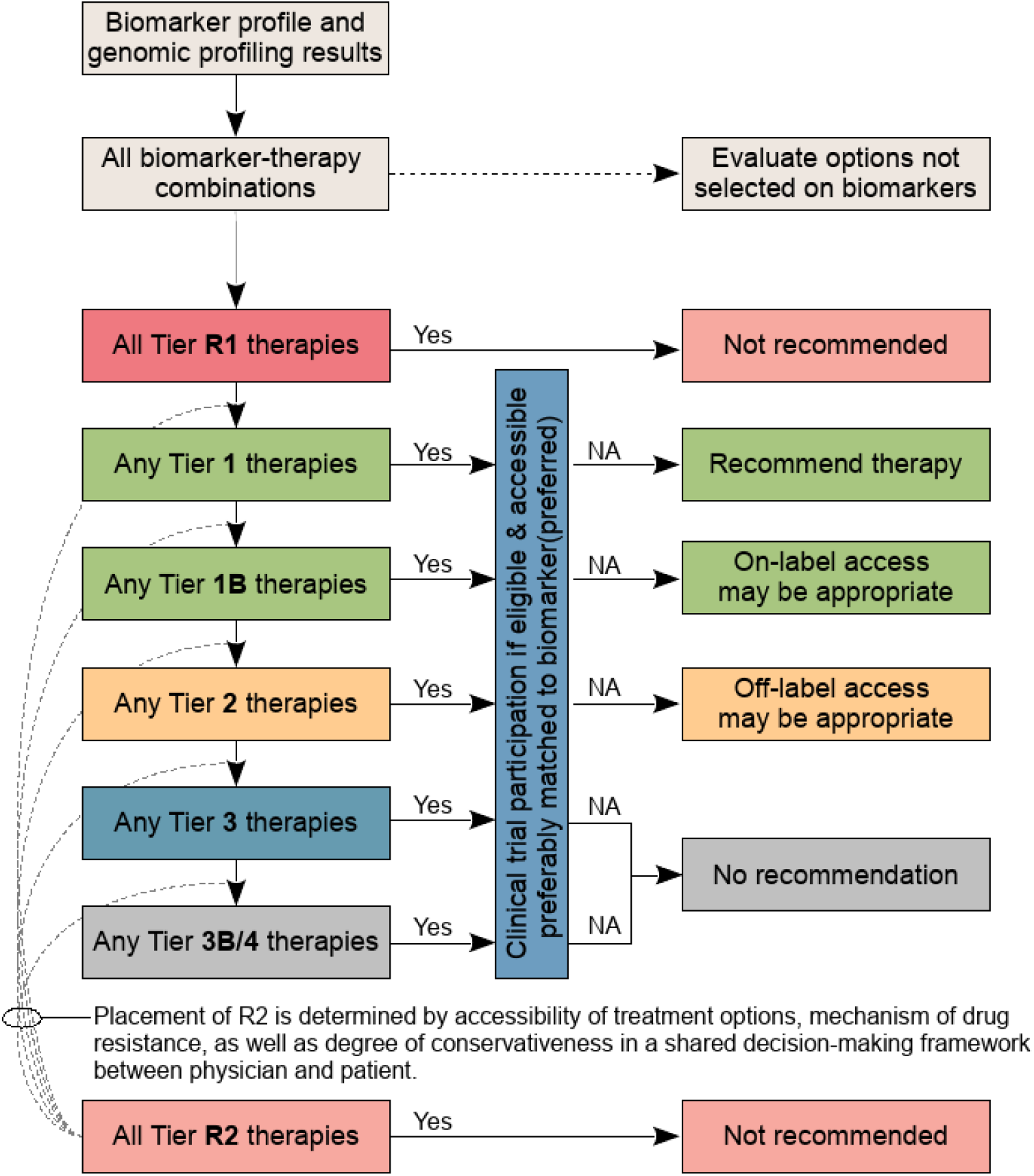
Proposed cascade decision algorithm to support treatment recommendation using biomarker driven therapies. In general, participation in a clinical trials should always be considered as best practice. T1 therapies are readily accessible and are thus recommended with exception of a concomitant, high-level resistance biomarker being present (i.e., Tier R1) or known treatment failure due to previous exposure, intolerance, or toxicity to another drug in the same therapeutic class. Off-label use of T2 therapies may be considered appropriate in selected circumstances. T3/4 therapies are not generally recommended outside clinical trial settings, given lack of compelling clinical data to support its use. In exceptional circumstances where treatment options are limited in rare cancer types, off-label access of lower-tier drugs may be appropriate. NA: therapy not available.

A remaining challenge is how best to integrate therapies with a R2 designation into the clinical decision-making process (Figure 3). These therapies are theoretically ineffective, although not yet proven in routine practice. Therefore, clinical discretion is needed to determine if the presence of the R2 resistance biomarker is sufficient to de-prioritise or override established treatment strategies, or perhaps to recommend such therapies within a clinical trials context. Broadly, the utility of R2 depends on the degree of conservativeness in patient management. The presence of a tier R2 biomarker should prompt discussion in the context of the patient’s condition, focusing on participation in relevant clinical trials and consider accessing alternative therapy.

### Challenges in assigning a tier to repurposed therapies

In patients with treatment-refractory disease, drug repurposing is not uncommon when a patient progresses through available standard-of-care therapies. When an exploitable, biologically rational molecular target is present (e.g., repurposing an anti-HER2 monoclonal antibody to treat a *HER2-*amplified tumour), oncologists may consider therapeutic regimens proven effective in another indication. Consistent with the LOE designation in OncoKB, we also assigned the repurposed therapies a *Tier 3B* designation – defined as treatments with demonstrable efficacy or antitumour activity (i.e., Tiers 1-3) in another cancer type when a biomarker is present. Despite its theoretical plausibility, however, disparate outcomes of treatments are often observed in patients treated with repurposed therapies in a histotype-dependent manner (e.g., distinct response patterns were seen in non-melanoma *BRAF* V600E cancers treated with vemurafenib [44]). As such, Tier 3B should be considered hypothesis-generating, such that patients are encouraged to enrol in clinical trials whenever possible. Alongside T4, therapies in this inferred tier have an important role in advancing drug development through expansion of indications into other disease entities.

## Discussion

Actionability is a complex concept. The results of molecular assays, by itself, only contribute to part of precision cancer care [28]. Issues such as implementation, targetability, and acceptability of treatment are integral to the decision-making process [22,25,31-32]. Here, we have created a pragmatic framework for therapeutic decision-making for oncologists faced with a molecular report. This framework broadly captures the important ‘real-world’ aspects of decision-making pertaining to drug treatments — local access, efficacy, and maturity with respect to its development cycle. Our literature framework builds on the therapy-focused concept proposed by the ESCAT criteria [33]. We believe TOPOGRAPH — a specific resource that implements this framework with ongoing efforts in curation — complements existing knowledge bases by filling the gap between biomarker interpretation and clinical decision-making.

While TOPOGRAPH consolidates the therapeutic aspects of molecular expertise, one cannot undermine the importance of the clinical context surrounding a patient and the multidisciplinary nature of oncology practice. For the latter, the importance of supplementing molecular expertise in cancer care through an MTB is increasingly recognised — both in improving the quality of decision-making and the rate of clinical trial participation [14-15,45-47]. A consistent decision-making framework is needed to deliver quality recommendations in this context. Reflecting on our experience over several years, we believe that TOPOGRAPH will also prove useful to MTBs by standardising recommendations with a higher degree of consistency. In the context of multidisciplinary decision making, specialised electronic resources have been shown to reduce decision variability in a complex decision-making setting [48].

TOPOGRAPH has several points of distinction from other resources that also catalogue biomarker-disease-therapy triplets. First, the evidence tiering system is tightly coupled with clearly defined literature criteria to reduce inter-reviewer variability. Second, the organisation of this compendium adapts a well-recognised LOE numbering system in broad categories to allow cross-database comparisons, albeit with some notable differences in the definitions. Third, an ongoing review strategy embedded within our MTB has significantly enriched the comprehensive and timely cataloguing of therapy-biomarker relationships, in addition to the active enrichment process of reviewing related biomarkers and therapies. Fourth, our curation strategy can accommodate the inclusion of emerging novel complex biomarkers (e.g., genome-wide assays for detecting homology recombinant defects [49-51]), not limited by single type of biomarkers (e.g., mutation). This “future-proofing” feature is important, given that rapid and continual emergence of both novel biomarkers and drugs is a perpetual feature of precision medicine [14,52]. Fifth, and perhaps most importantly, our evidence tiers aim to facilitate evaluation of strength-of-recommendation for each therapy. This approach complements those knowledge bases focused primarily on variant interpretation rather than actionability of individual therapeutics (Figure 4). Together, these features enable TOPOGRAPH as a decision support tool for sieving through available drug options based on molecular biomarkers in routine clinical practice.

**Figure 4.**
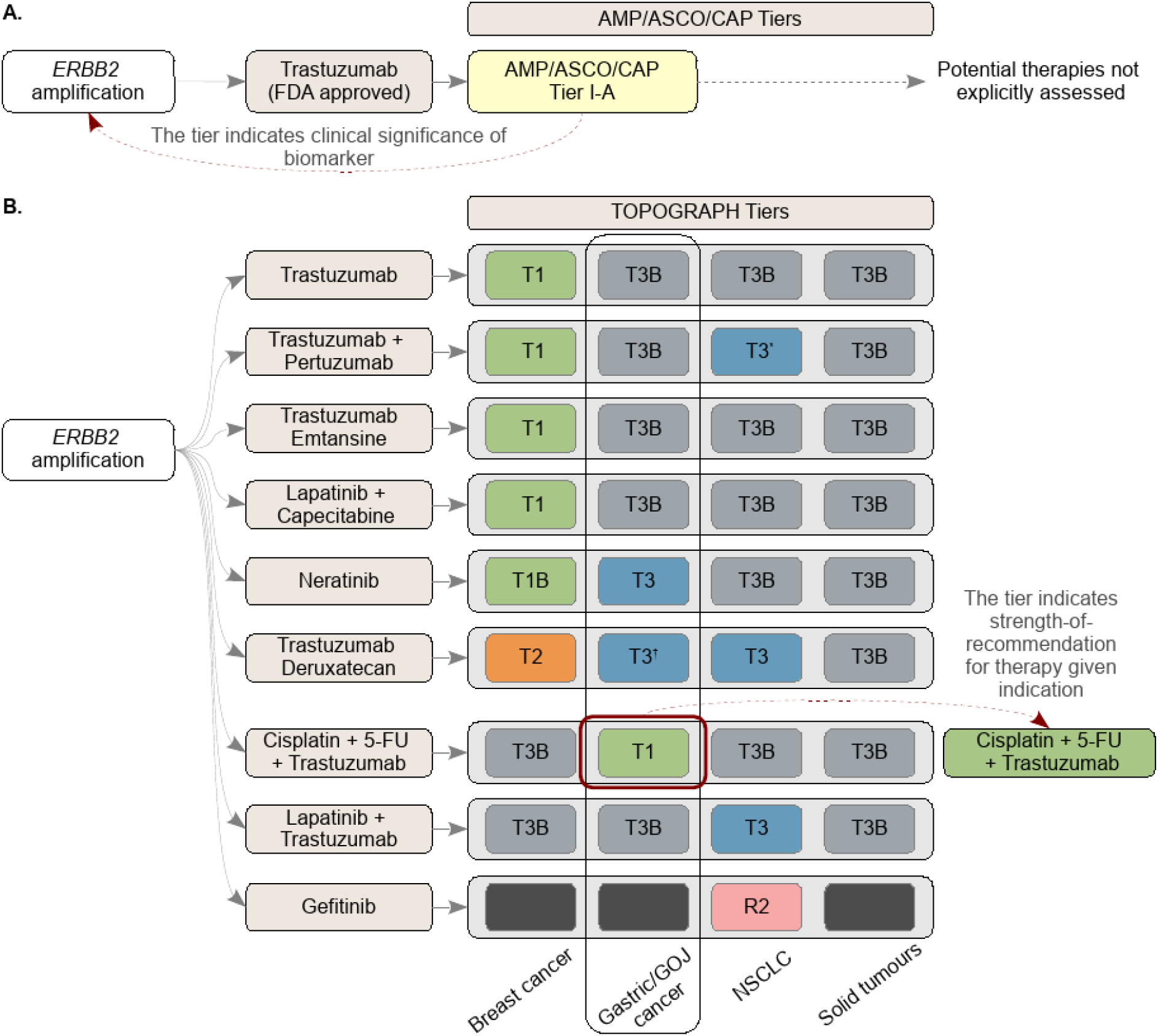
A comparison of therapy- and biomarker-focused tiering approach. A schematic diagram comparing the two different tiering approaches. (A) In a biomarker-focused approach (e.g., AMP/ASCO/CAP system), a tier is assigned to a molecular alteration to indicate its clinical significance; complete list of therapies is not explicitly assessed. (B) In contrast, TOPOGRAPH – which uses a therapy-focused approach to review literature – assigns a tier to each therapy (instead of a biomarker) to indicate the strength-of-recommendation based on available evidence. This approach identifies and ranks potentially actionable treatment options with respect to the clinicopathologic contexts (e.g., cancer type). Abbreviations: AMP: Association for Molecular Pathology; ASCO: American Society for Clinical Oncology; CAP: College of American Pathologists; FDA: U.S. Food and Drug Administration; FU: Fluorouracil; GOJ: Gastro-oesophageal junction; NSCLC: Non-small-cell lung cancer.

We also aimed (1) to support localisation of precision oncology knowledge, and (2) to facilitate biomarker-based design of clinical trials. While this paper describes the use of TOPOGRAPH in the Australian context, our approach can be extended to other jurisdiction-specific guidelines through re-tiering of standard-of-care treatments (T1/2). From a global oncology perspective, comparing T1/2 therapies between countries may help identify differences in equity of access, highlighting the disparity in drug utilisation compared to scientific advances in cancer therapeutics. Given that patients’ access to treatment varies considerably across health systems, many treatments are only accessible through clinical trials which may or may not be accessible regionally. In addition, there is a potential role for TOPOGRAPH to support translational research, through informing the design of new correlative studies to explore more precise biomarkers in selecting patients for targeted therapies.

Several limitations of our work are also noted and the need for ongoing refinement is actively being investigated. First, as precision oncology is constantly evolving, TOPOGRAPH and other similar knowledge bases require continual curation to remain relevant with timely distribution of knowledge; automated information retrieval and categorisation may accelerate this ongoing effort with more prompt updates [53,54]. Second, for any given patient, this compendium must be used in the context of other non-targeted and established therapy options. TOPOGRAPH also relies on co-dependent developments in molecular pathology and bioinformatics, including novel feature identification and assignment of variant pathogenicity, which form the basis for therapy selection. Third, the present definition of T4 and R2 tiers include a broad range of clinical and preclinical evidence about a therapy. Negative data tend to be poorly reported in published literature, which disproportionately affects the R category. Further refinements in defining these tiers can facilitate trial design, as can efforts to increase reporting of both negative as well as positive data from trials. Fourth, no knowledge base currently quantitatively considers the magnitude-of-benefit of therapies, which is both drug and context dependent. Combining interpretation of targetable cancer biology in the context of established clinical care pathways will be an important part of the evolution of decision-support systems. Fifth, continuous monitoring of post-marketing evidence of conditionally approved drugs is important to ensure that the surrogate endpoint about efficacy of a drug can be affirmed [55]. An indication of a therapy may also be withdrawn if evidence generated from subsequent post-approval studies do not support its projected effectiveness [56]. Sixth, the rapid evolution of knowledge in precision oncology prohibits repeated systematic review (SR) using traditional publication methods. While it is practically infeasible for a curated knowledge base to scrutinise results of all clinical trials to the same extent as in SR, its breadth allows relevant literature to be readily retrieved for synthesis by oncologists at the bedside. However, the use of a tiered knowledge base to make recommendations remains untested, and research is needed to examine if this decision-making strategy will lead to clinical benefits in prospective studies.

In summary, we have developed a framework for collating information and evidence relevant to therapy decision-making in precision oncology, and it has facilitated the curation of a jurisdiction-specific precision oncology resource to aid decision-making. The proposed decision algorithm, together with the curated knowledge base, warrants further examination of its utility with respect to treatment recommendations supported by an electronic compendium.

## Methods

### Database design

The TOPOGRAPH database curated triplets of information comprising predictive biomarker, cancer type, and therapy (biomarker-disease-therapy triplet, thereafter triplets). *Biomarkers* consist of gene alterations (including simple mutations, copy number variations, and structural variants), variations in protein expression (e.g., over-expression or loss of expression), or synthetic molecular characteristics (e.g., tumour mutational burden, TMB) associated with an outcome measure for a particular therapy. The *disease* category broadly defines cancer types listed in the literature or indicated for a therapy. *Therapies* comprise approved or experimental drugs, and their combinations, represented using standardized International Non-proprietary Names. References to literature supporting the curation of each triplet were recorded.

### Data sources

The therapy list was compiled by identifying all cancer drug lists (and combinations) from historical drug approvals by regulatory authorities (TGA Australia and U.S. FDA); the registered indications were extracted. For drugs registered in Australia, the corresponding status of reimbursement was extracted from Australian Pharmaceutical Benefit Schedule (PBS). Therapies listed in National Comprehensive Cancer Network (NCCN) guidelines and therapies curated in three knowledge databases were also reviewed [19,22-23,57]. For a given cancer type, the evidence for differential efficacy or therapy response in the presence of a biomarker were manually appraised from published journal articles and abstracts from MEDLINE and major international oncology congresses.

The initial curation process also included a merger of three local knowledge sources, including a local knowledge base generated by the molecular tumour board (MTB) of a national precision oncology program (The Molecular Screening and Therapeutics study [5]), and two independently curated databases by two oncologists (FL & ST). Data were accessed and reviewed between April and November 2020.

### Standardisation of biomarker nomenclature

Catalogued predictive biomarkers included genes and alterations (e.g., *BRAF* V600E, *ALK* fusions); protein expressions (e.g., PD-L1 expression of tumour cells as determined by immunohistochemistry assays) and other novel genomically derived biomarkers (e.g., TMB, homologous recombination deficiency scores). Alterations of biomarkers were abstracted to remove proprietary information. Gene names were standardised to HUGO Gene Nomenclature Committee (HGNC) symbols [58]; Human Genome Variation Society (HGVS) Sequence Variant Nomenclature was used to describe gene mutations [59], defaulting to protein sequences. Expression of protein was designated to the corresponding HGNC gene symbols wherever possible.

### Strategy for literature appraisal

After initial curation, literature appraisal was conducted by two medical oncologists (FL and ST) using the following process: literature was reviewed if a therapy was (1) curated in one of the publicly available knowledge bases; (2) being studied in a clinical trial with explicit mention of using biomarkers for stratification or therapy selection, as documented in ClinicalTrials.gov or disseminated in publications; (3) examined as a potential biomarker of response or efficacy in exploratory analysis in a clinical trial or retrospective studies; (4) examined in preclinical studies regarding antitumour activities; or (5) mentioned in a published review article where a potential therapy (or therapy class) was suggested to have association with a predictive biomarker.

Conversely, literature was excluded if no biomarker was specified or only implied in certain cancer types (e.g., up-regulation of PI3K/mTOR pathway in renal cell carcinoma). As described above, *active exploration* and appraisal were further conducted on each curated triplet to expand the curation of related biomarkers, therapies, and cancer types (Figure 1).

### Evidence tiering system

To enable comparison of content between TOPOGRAPH and other resources, we constructed the evidence tiering system based on the numeral designations of level-of-evidence (LOE) developed by OncoKB [22]. This system was selected on the basis of the LOE mostly oriented to clinical recommendations, compared to other systems. The ESCAT system was not adopted due to insufficient ability to represent therapy resistance [33]. Similarly, the Association for Molecular Pathology, American Society of Clinical Oncology, and College of American Pathologists (AMP/ASCO/CAP) criteria were not used as it primarily focuses on variant interpretation [26]. Specific literature criteria were then developed to systematically grade literature that supports the tier assignment of each triplet (Table 1).

### Data availability

The data and a web resource are available online at https://topograph.info/.

## Data Availability

Data is available online at https://topograph.info/.

## Acknowledgements

The authors declare no conflict of interest. The Australian Genomic Cancer Medicine Program (Omico Ltd) is funded by the Office for Health and Medical Research, State of New South Wales and Australian Federal Government, and acknowledges support from Roche for contributing to the Molecular Screening and Therapeutic (MoST) trial. The authors thank Lucille Sebastian for commenting on an earlier version of the manuscript. FL acknowledges support from Philanthropic funds of the Wolf family for early part of this work. DT is supported by the National Health and Medical Research Council (NHMRC) Principal Research Fellowship.

## Author Contributions

Database design and concepts: F.P.L., S.T., J.P.G., J.S., J.D., D.M.T.; Content curation: F.P.L., S.T., J.P.G., S.O., M.K., D.M.T. Data analysis and maintenance: F.P.L. Critical appraisal, manuscript writing, and correction: all authors.

