## Supplemental text for "Criteria-based curation of a therapy-focused compendium to support treatment recommendations in precision oncology"

### Table S1: An excerpt from TOPOGRAPH showing differential tiering of tumour mutational burden as a predictive biomarker for immune checkpoint inhibitors.

| Tier | Biomarker | Alteration | Cancer type | Therapy | Comments |
|---|---|---|---|---|---|
| 2 | TMB | High | Cervical cancer<br>Endometrial cancer<br>Neuroendocrine tumour<br>Salivary gland carcinoma<br>Small-cell lung cancer<br>Thyroid cancer<br>Vulvar cancer | Pembrolizumab | Not TGA approved. FDA approved. In Phase 2 KEYNOTE-158 trial, the following cancer types have higher response rate in TMB high group compared to non-TMB-H groups. |
| 2 | TMB | High | Non-small cell lung cancer | Nivolumab + Ipilimumab | Not TGA approved. Phase 3 CHECKMATE 227. Prespecified, prospective TMB cutoff of 10mut/MB in 57% of cohort. Median PFS: 7.2 vs 5.5 months. Quantitative TMB is not correlated with TMB. |
| 3 | TMB | High | Non-small cell lung cancer | Durvalumab + Tremelimumab | Pre-planned exploratory biomarker analysis from the Phase 3 MYSTIC trial. |
| 3 | TMB | High | Solid tumours except Colorectal Cancer | Pembrolizumab | Overall Tier 3 based on phase 2 KEYNOTE-158 trial, forming the basis of FDA pan-cancer approval noted. In overall cohort, TMB-H (v non-TMB-H) was noted to have an objective response rate of 29% (versus 6%). |
| 4 | TMB | High | Colorectal Cancer | Pembrolizumab | Microsatellite-stable colorectal cancers were not included/reported in KEYNOTE-158. |
| 4 | TMB | High | Non-small cell lung cancer | Nivolumab | Exploratory analysis from Checkmate 026. |
| 4 | TMB | High | Gastric cancer | Toripalimab | Exploratory analysis showed ORR 33% v 6% at TMB threshold of 12/MB |

Abbreviations TMB: Tumour Mutational Burden.
